# Designathons in Health Research: A Global Systematic Review

**DOI:** 10.1101/2023.07.17.23292758

**Authors:** Warittha Tieosapjaroen, Elizabeth Chen, Tiarney Ritchwood, Chunyan Li, Jamie L. Conklin, Abdulhammed Babatunde, Arturo Ongkeko, Ucheoma Nwaozuru, Joseph D. Tucker, Jason J. Ong

## Abstract

**Background:** A designathon is a three-stage participatory activity informed by design thinking and rapid prototyping that includes preparation with end-users, an intensive period of collaborative teamwork, and evaluation of solutions by topic experts or community partners. A few previous systematic reviews have focused on the use of designathons in health. This study synthesised how designathons were organised and implementation-related factors to address health challenges.

**Methods:** We searched Cochrane Library, Embase, PubMed, Scopus, and the ClinicalTrials.gov registry for peer-reviewed articles until November 29, 2022. The systemic review was registered in PROSPERO (CRD42023389685).

**Results:** In total, 4,947 citations were identified, with 38 studies included in this review. Most studies were from high-income countries (26, 68%). The median number of participating teams was eight (IQR 5, 15), and the duration of the intensive collaboration phase ranged from three hours to seven days. The final products (i.e., ideas and prototypes) related to four themes: mobile applications, educational programs, medical devices, and other prototypes. Common evaluation criteria were feasibility, innovation or creativity, and impact. The most common facilitators were including diverse participants and having high-quality mentorship. . The most common barriers related to planning and implementing the designathon, and engaging diverse participants to participate. There were limited data on required resources and further implementation of solutions after designathons and no data on cost-effectiveness.

**Conclusion:** Designathons are a promising tool for fostering innovative and person-centred solutions to address health challenges. Given its adaptability in terms of budget, mode of delivery, and involvement of diverse participants including end-users, designathons can be implemented in a wide range of contexts to address various health issues.

## Introduction

A designathon is a time-based three-stage participatory activity informed by design thinking and rapid prototyping that includes preparation with end-users, an intensive period of collaborative teamwork (typically from a few hours to a few days), and follow-up activities for implementation and research (including the evaluation of solutions by topic experts or community partners). This approach is a type of crowdsourcing activity where ‘bottoms-up’ solutions are generated by a large group or ‘crowd’ instead of relying on internal experts (the ‘top-down’ approach).^1^ Designathons directly engage multiple stakeholders, including end-users, increases public awareness of specific health challenges (e.g., multiple cultural settings) and foster a sense of community empowerment and collective responsibility, leading to more sustainable and impactful outcomes ^4^ Designathons are related to different methods, including hackathons, co-creation sessions, and design sprints. Designathons are similar to those listed above in that they try to solve a problem^2^ and offer a specific, structured process. Designathons differ from those listed above in that they explicitly integrate design thinking or human-centred design principles into their processes.^3^

Designathons are increasingly used to engage end-user communities in multiple settings to generate socially innovative solutions to health challenges.^5, 6^ For instance, more than 26,000 individuals attended an online designathon hosted by Germany’s government to develop solutions to address the Covid-19 pandemic.^7^ Additionally, designathons have been used by The National Academies of Sciences, Engineering, and Medicine,^8^ the World Health Organization (WHO),^9^ and the United States National Institute of Health .^10^

Given that few reviews have analyzed all three phases of designathons for health,^11–13^ our systematic review is unique because we used a complementary designathon to identify best practices and inform a WHO/TDR practical guide. We aim to synthesize data from designathons used in health research in the peer-reviewed literature to understand how they are implemented, their effectiveness (i.e., engagement, outputs and implementation), required resources, benefits, drawbacks, facilitators and barriers.

## Methods

We followed guidance from the Cochrane Collaboration to conduct the systematic review and reported our findings according to the Preferred Reporting Items for Systematic Reviews and Meta-Analyses (PRISMA) 2020 statement.^14^ We registered the protocol in PROSPERO (CRD42023389685).

### Eligibility Criteria

Studies were included if they met the following eligibility criteria: 1) contained primary data regarding designathons for health (i.e., time-limited event), involved a range of stakeholders in working in teams to design solutions to a specific problem, solutions were judged or evaluated by a panel, the solutions might be implemented or tested after the event); 2) focused on health outcomes related to the attainment of either physical, mental and/or social well-being of human beings or the healthcare system and/or policy; and 3) the event mentioned explicitly design thinking or human-centred design, or included participant activities focused on design thinking steps including empathize, define, ideate, prototype, and evaluation. Studies were excluded if they were duplicates of other studies.

### Search Strategy and Study Selection

A health sciences librarian searched the following four databases from their inception to November 29, 2022: the Cochrane Library, Embase (Elsevier), PubMed, and Scopus (Elsevier). The ClinicalTrials.gov registry was also searched. The searches included a combination of subject headings and keywords for the main concept of designathons. Four separate approaches were combined with OR: terms for designathons or agile methodology; user-centred design or rapid prototyping events; hackathons, makeathons, and other similar events; and community participatory ideation events. The Scopus search had an additional search string related to health and well-being since the database includes disciplines beyond health and medicine. Conference abstracts were filtered from the Embase search. No other filters or restrictions were placed on the searches. The complete and reproducible search strategy can be found in Table S1.

The librarian (JC) placed all references into Endnote X9 (Philadelphia, Pennsylvania, USA) and removed duplicates. We used a two-stage automation approach to prioritize the screening process. First, the team independently screened 300 references to identify seed studies as training data for supervised clustering through ICF’s Document Classification and Topic Extraction Resource (DoCTER) (https://www.icf-docter.com/). Each reference was given an ensemble score ranging from 0 (deemed irrelevant) to 6 (deemed relevant in all six models). Those with a score of four or higher were imported into Covidence systematic review software (Veritas Health Innovation, Melbourne, Australia, available at www.covidence.org) for screening. Each abstract was screened for eligibility criteria by two researchers independently (WT, TR, EC, CL, AO, AB, UN), with the third reviewer (JJO) resolving any conflicts that arose.

For the second screening stage, studies moved to full-text review through the first stage were used as training data for machine learning, also through DoCTER. Each unscreened study was given a probability score ranging from 0 to 1, with a higher score indicating a higher relevance ranking. Each title and abstract were again screened by two researchers in Covidence until relevance dropped off, which was at a probability score of 0.4. As in the first stage, each full-text article was screened for eligibility criteria by two researchers independently (TR, EC, CL, AO, AB, UN), with the third reviewer (WT) resolving any conflicts that arose. We also screened additional potentially relevant peer-reviewed studies we received from the complementary open call for examples of designathons. This was conducted between January 16^th^, 2023 and March 7^th^, 2023, and received 27 submissions.

We used descriptive statistics (including percentages for categorical variables and median and mean for continuous variables) to summarize designathon characteristics, such as region of study, designathon participants, and final products of designathons. We conducted a content analysis to describe shared themes related to the outputs, barriers, facilitators, strengths and limitations to implementing specific designathons, along with more generally utilizing designathon approaches towards collecting data.

### Quality Assessment

We assessed the risk of bias using a checklist adapted from Joanna Briggs Institute Critical Appraisal tools.^15^ Risk of bias was independently examined by two reviewers (WT and EC), and any discrepancy was resolved by a third reviewer (JJO). However, this did not influence the inclusion of the studies in this review (Table S2).

### Results

Overall, 4,846 citations were identified via electronic database searching, with 31 publications included in this review. One-hundred-and-twenty-three publications were retrieved from additional searches (i.e., citation searching and the complementary open call), with seven publications included in this review. In total, we included 38 studies in this review.^16–22^ The PRISMA flowchart is presented in Figure S1. We included 26 publications (68%) from high-income countries, nine (24%) from middle-income countries, two (5%) global publications and one publication (3%) with an unclear location. Thirty-two publications (84%) presented data on in-person designathons, and 30 (79%) used ‘hackathon’ to describe designathons. Twenty-five publications (66%) were case reports. Nine (24%) were mixed-method studies, and four (11%) were qualitative studies. No randomised controlled trials were included. The final product from designathons were ideas (N=18, 47%) and prototypes (N=18, 47%) (Table 1).

**Table 1.**
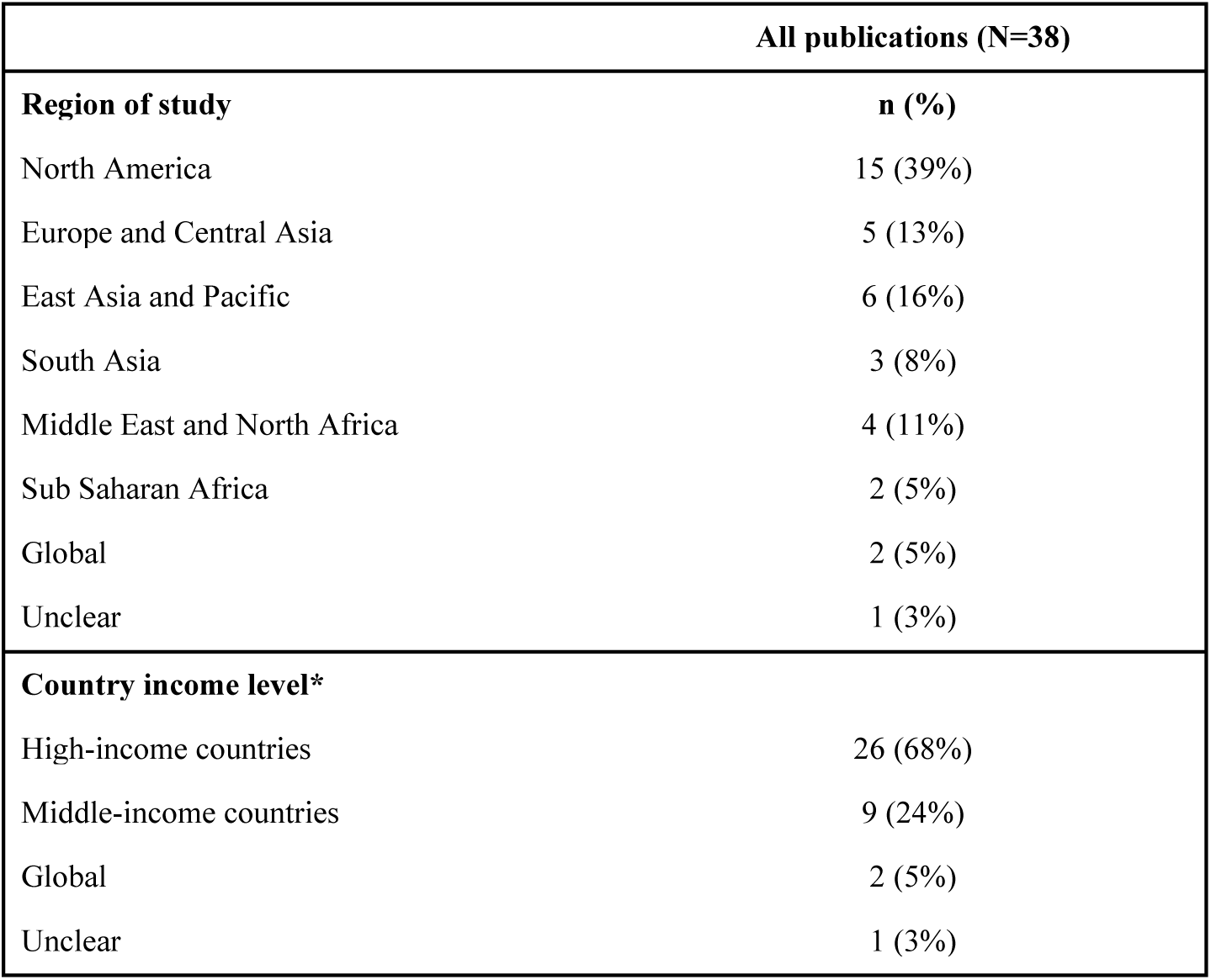
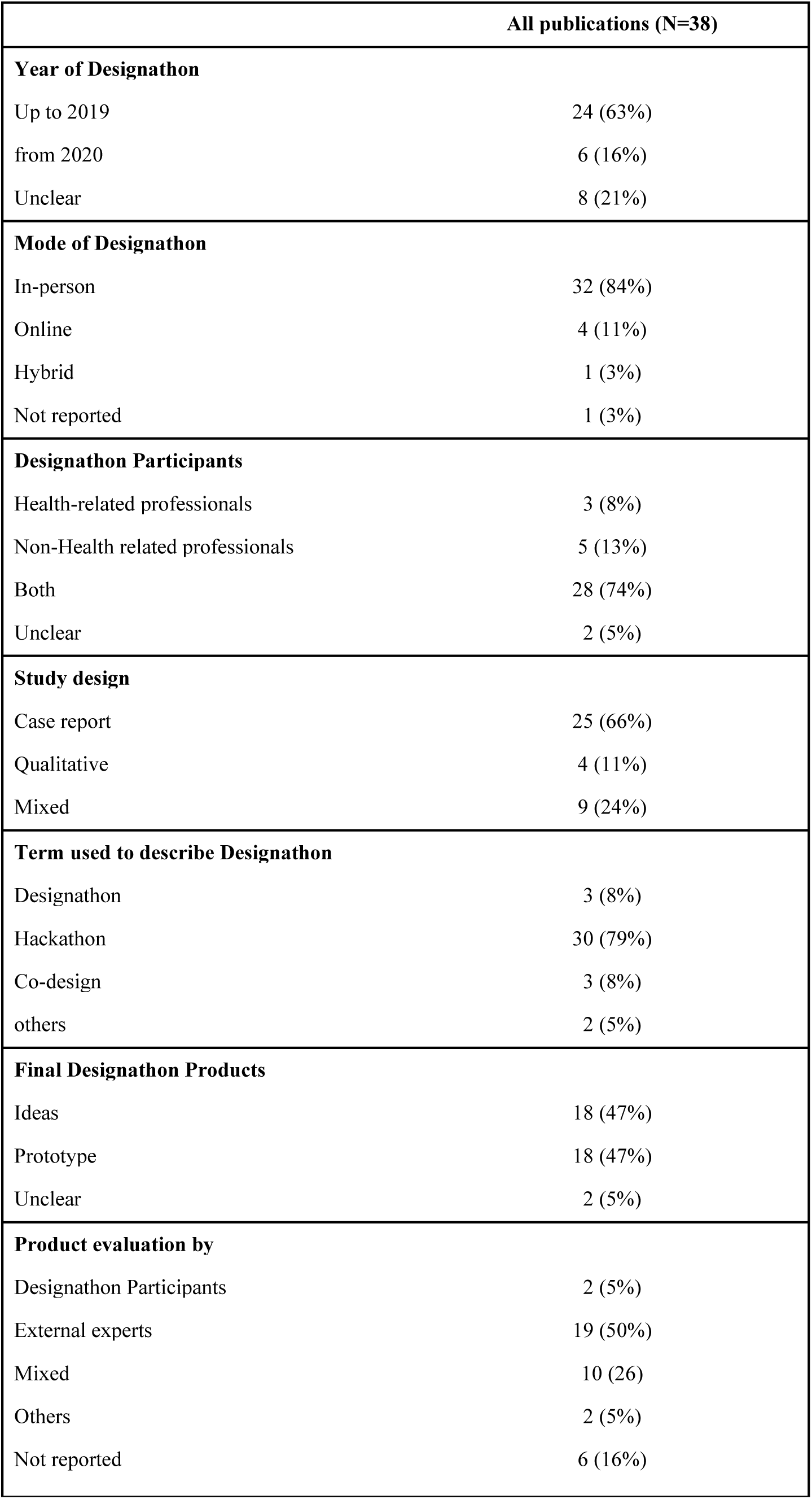
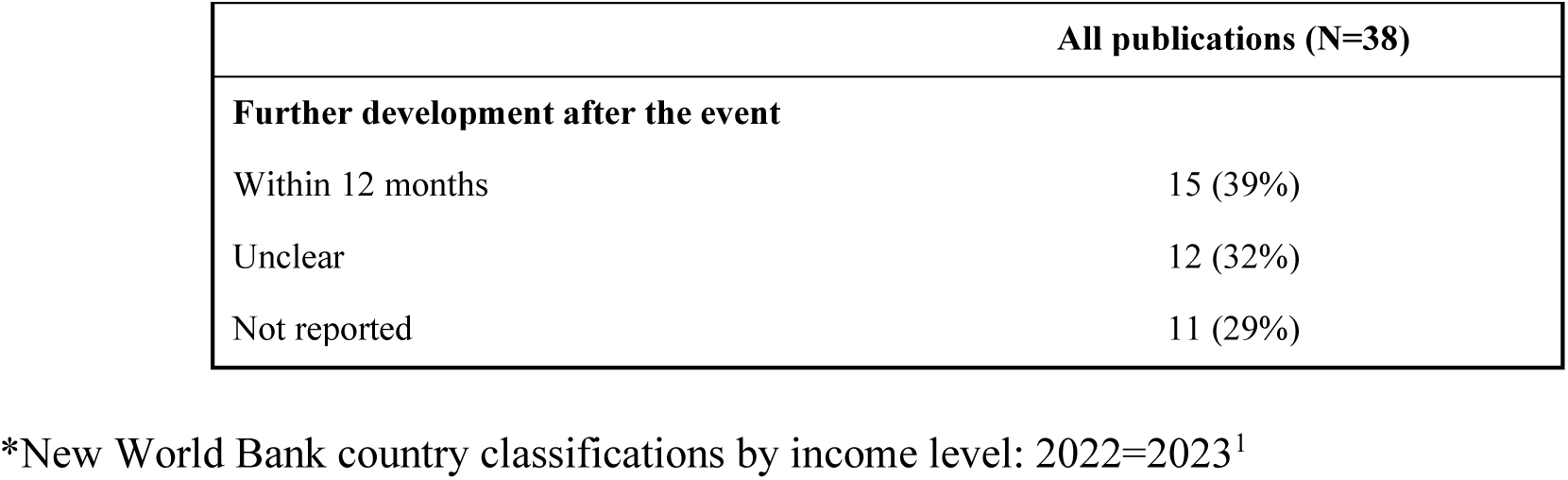
Overview of the studies included.

### Engagement

Thirty-seven studies (97%) reported the number of designathon participants.^3, 5, 6, 16, 18–50^ The median number of individuals participating in designathons was 60 (IQR 34.5, 96, range 6-434), and the median number of teams was 8 (IQR 5, 15, range 2-40). Thirty-three studies (87%) reported the duration of the intensive collaboration part, ranging from 3 hours to 7 days (Table 2).^3, 5, 6, 16, 18–23, 26–28, 30–33, 35–45, 47–51^

**Table 2.**
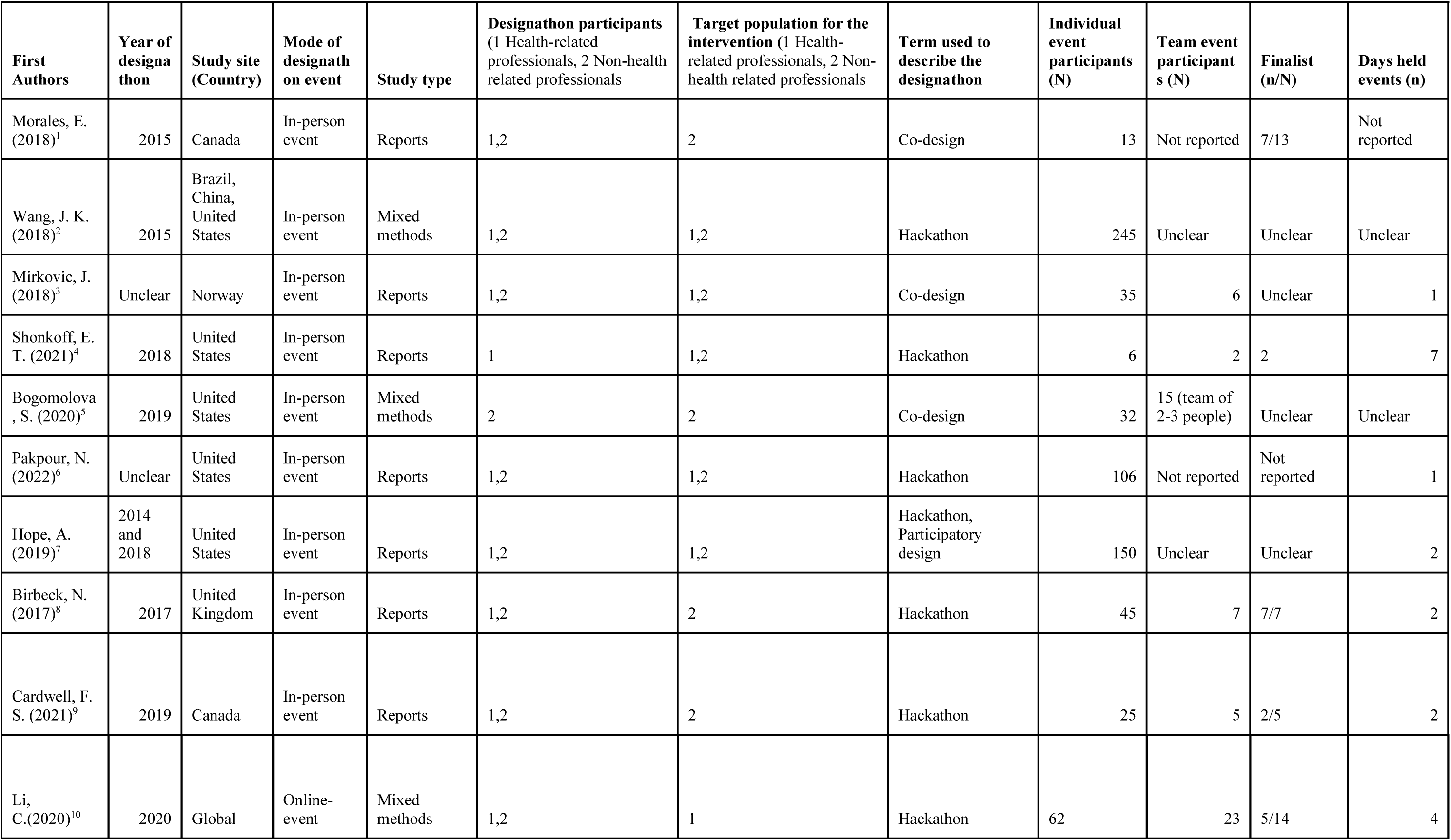
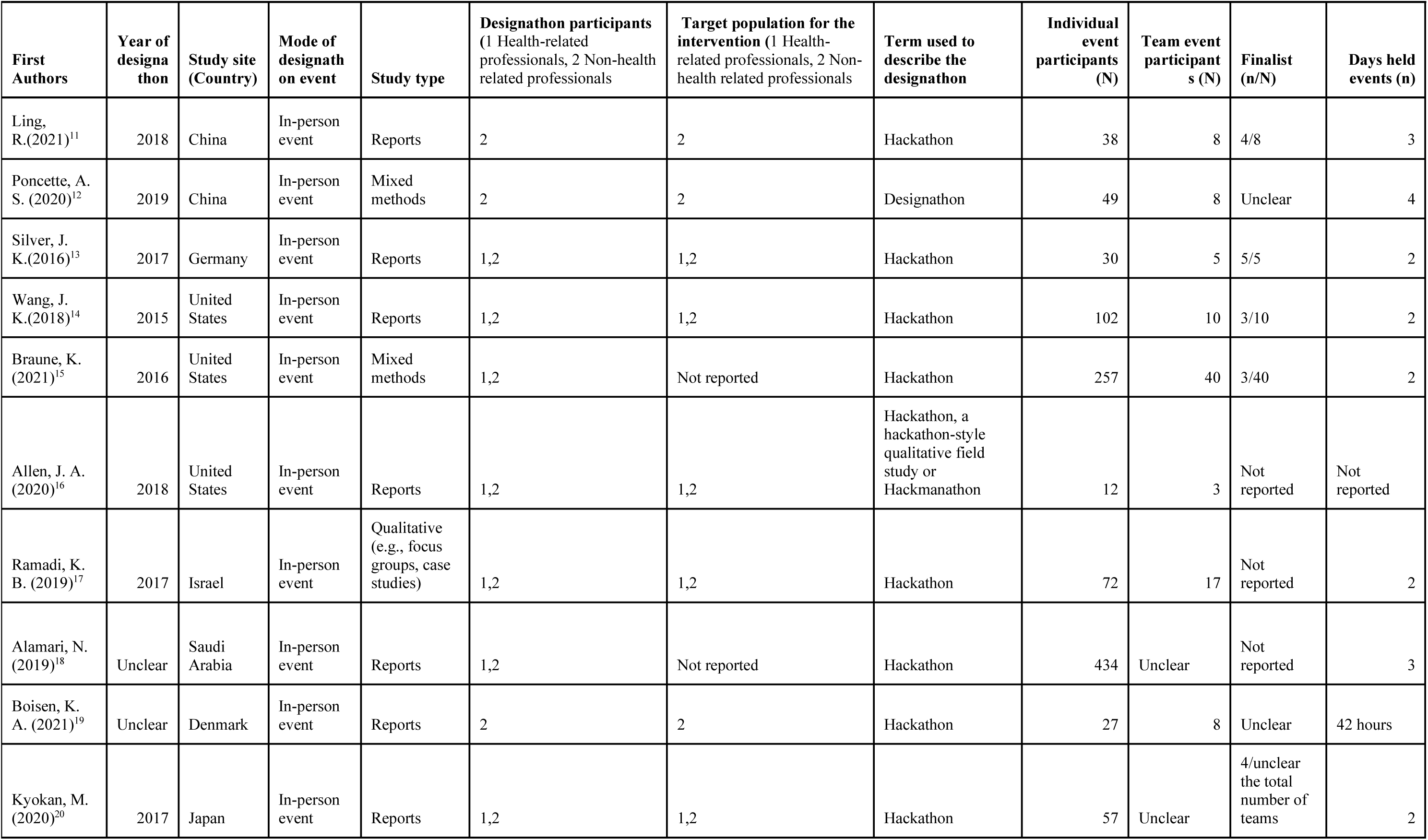
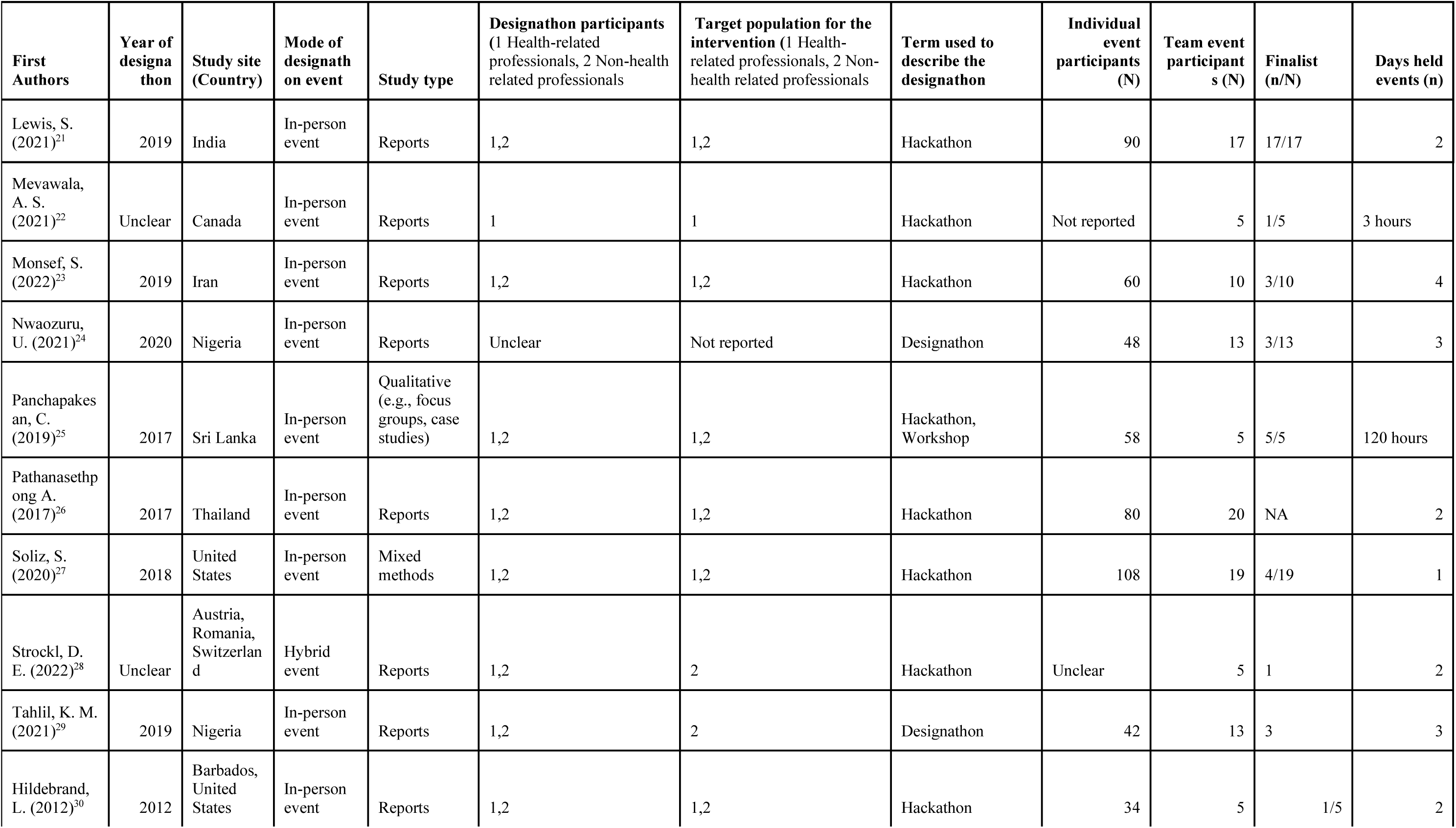
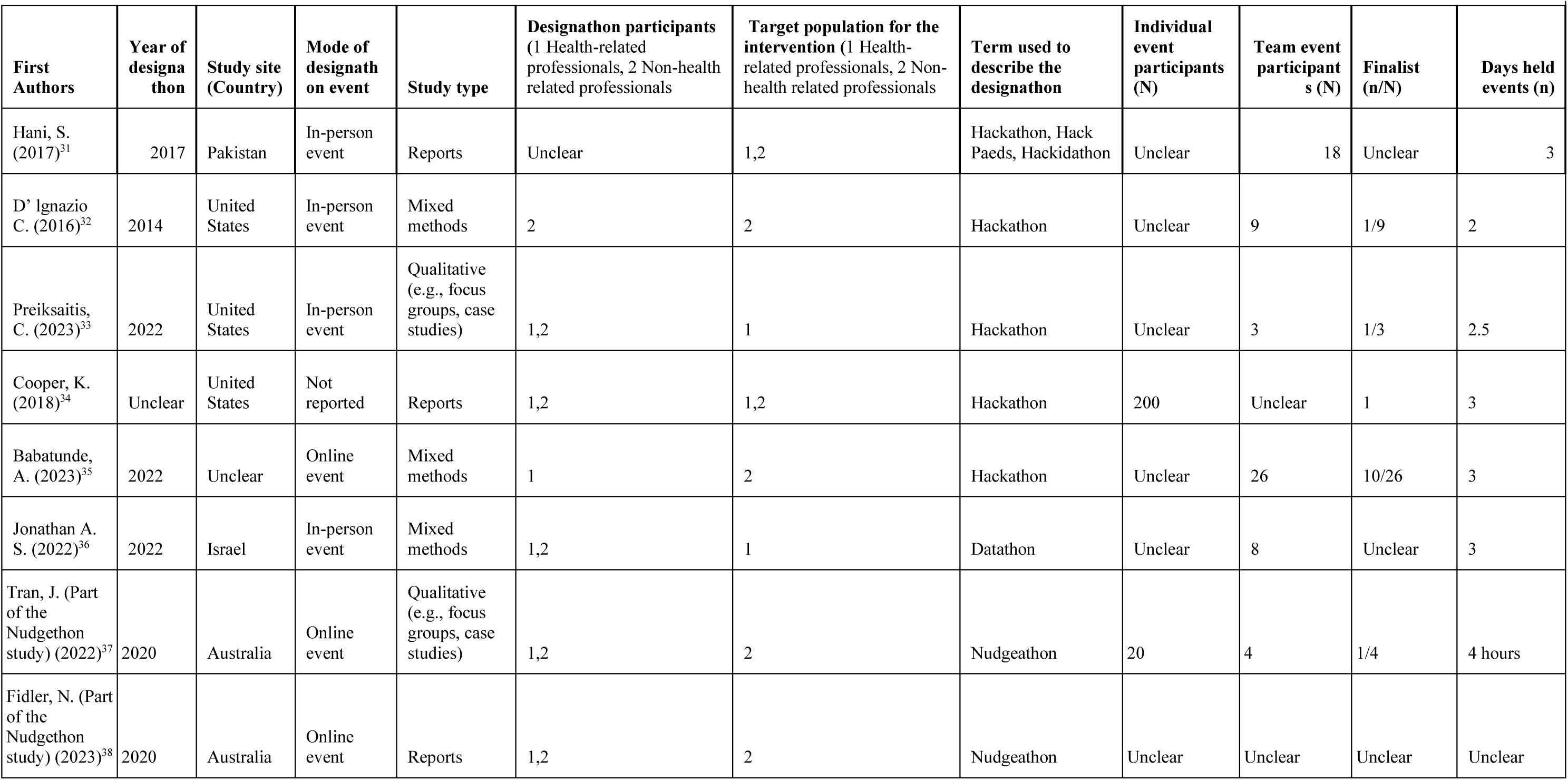
Characteristics of publications included (N=38)

### Resources

Only two studies (5%) reported data on the cost of organising a designathon.^33, 39^ A 2-day designathon organised in India to develop solutions for cancer care reported that each participant cost around 16 British Pounds.^39^ Meanwhile, A two-day hackathon in the U.S. aimed at generating solutions for unmet clinical needs cost approximately US$40,000. No cost-effectiveness studies were found.^33^

### Outputs

Twenty-five studies (66%) reported the number of final products (i.e. ideas or prototypes) generated during the event.^3, 5, 6, 18, 19, 21, 23–31, 33, 34, 38–42, 45, 47^ The median number of the final product was 8.5 (IQR 5, 13, range 2-40). The final products reported from thirty-two studies (84%) emerged as four themes: 1) educational programs (n=8, 21%), such as wellness and healthy choice programs;^16, 17, 23, 26, 27, 29, 44, 46^ 2) mobile phone applications (n=7, 18%), such as management of opioids and support mental health;^20, 21, 28, 30, 31, 40, 41^ 3) medical devices (n=4, 11%), such as breast pumps and humanitarian surgical care;^3, 18, 25, 35^ and 4) mixed interventions (n=11, 29%) (Table 3).^5, 6, 19, 24, 32–34, 36, 39, 42, 48^ Regarding evaluation, 20 studies (53%) reported evaluation criteria, with the common criteria being feasibility, innovation or creativity, and impact.^3, 5, 6, 19, 21, 24–26, 28, 30, 31, 33, 36, 37, 39, 40, 42, 44, 49, 51^ Eight studies (21%) reported using other methods, such as feedback from the judge panel and audience and votes, to evaluate the product generated during the designathon (Table 3).^16–18, 20, 29, 30, 45, 46^ The judge panels included designathon participants (N=2, 5%),^29, 30^ external expert (N=18, 47%)^5, 18, 19, 21, 22, 28, 32–34, 36–38, 42, 43, 45, 48–50^ and more than one professions (N=11, 29%) (Table 3).^6, 23, 25–27, 31, 39, 40, 44, 46, 47^

**Table 3.**
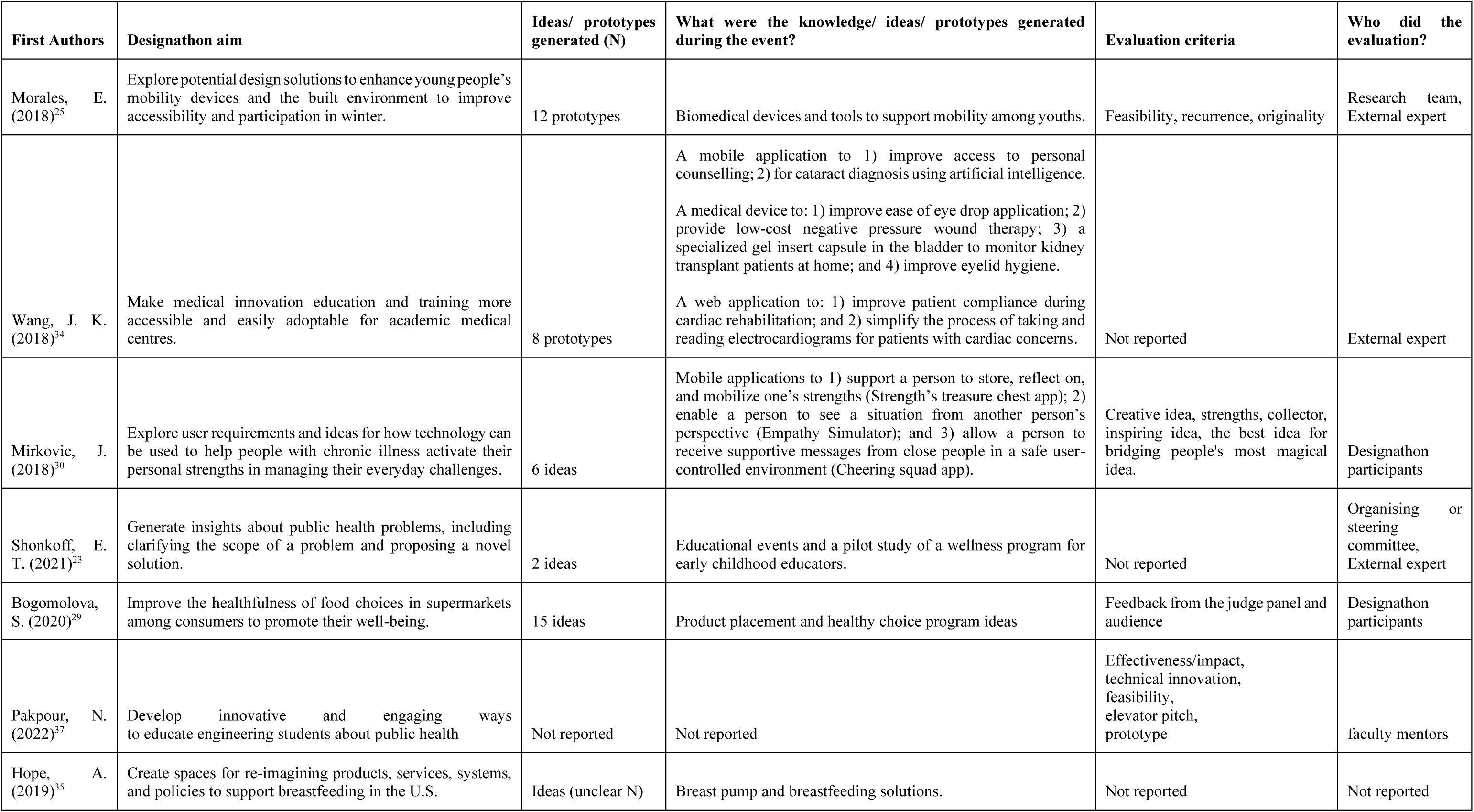
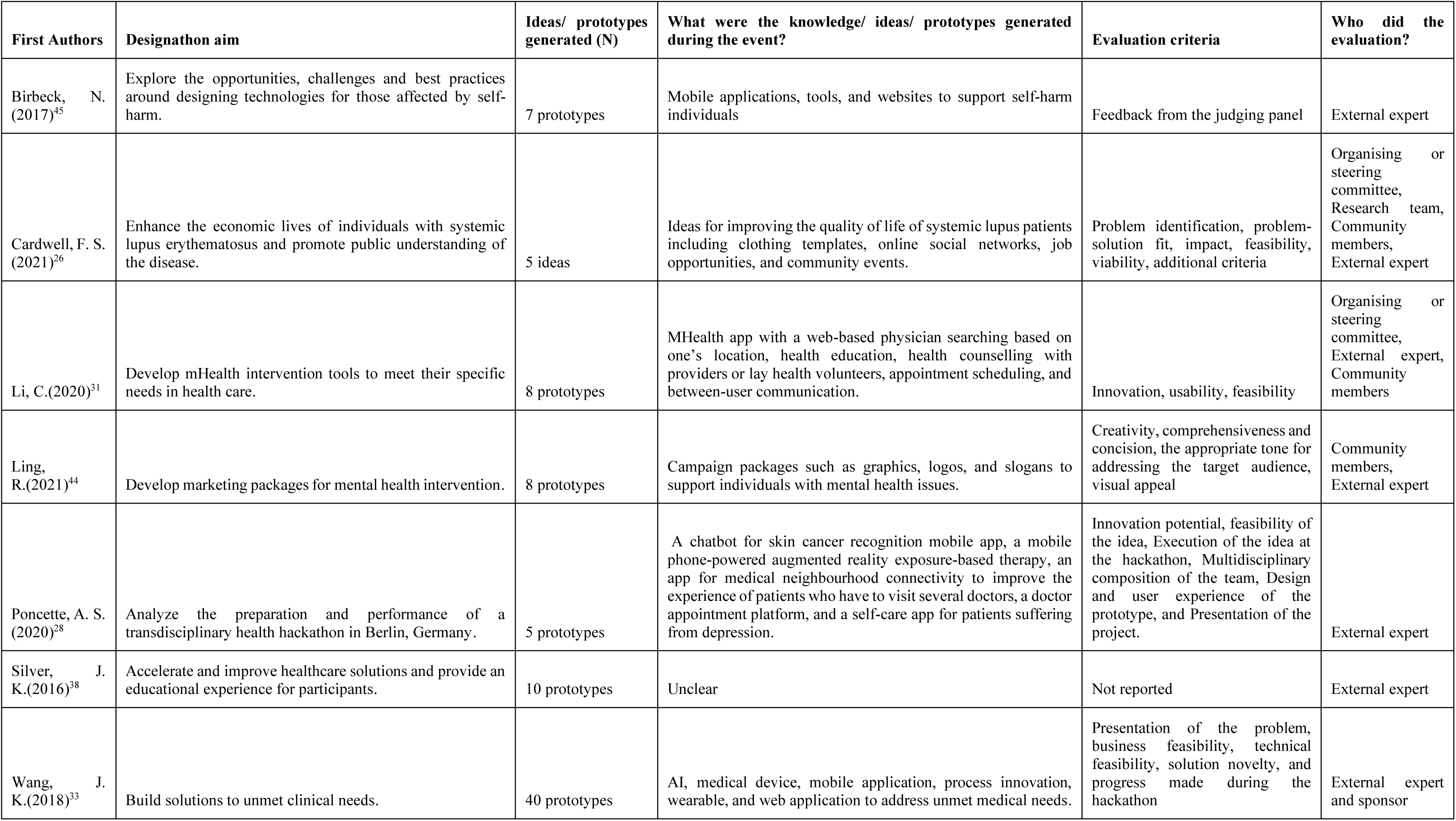
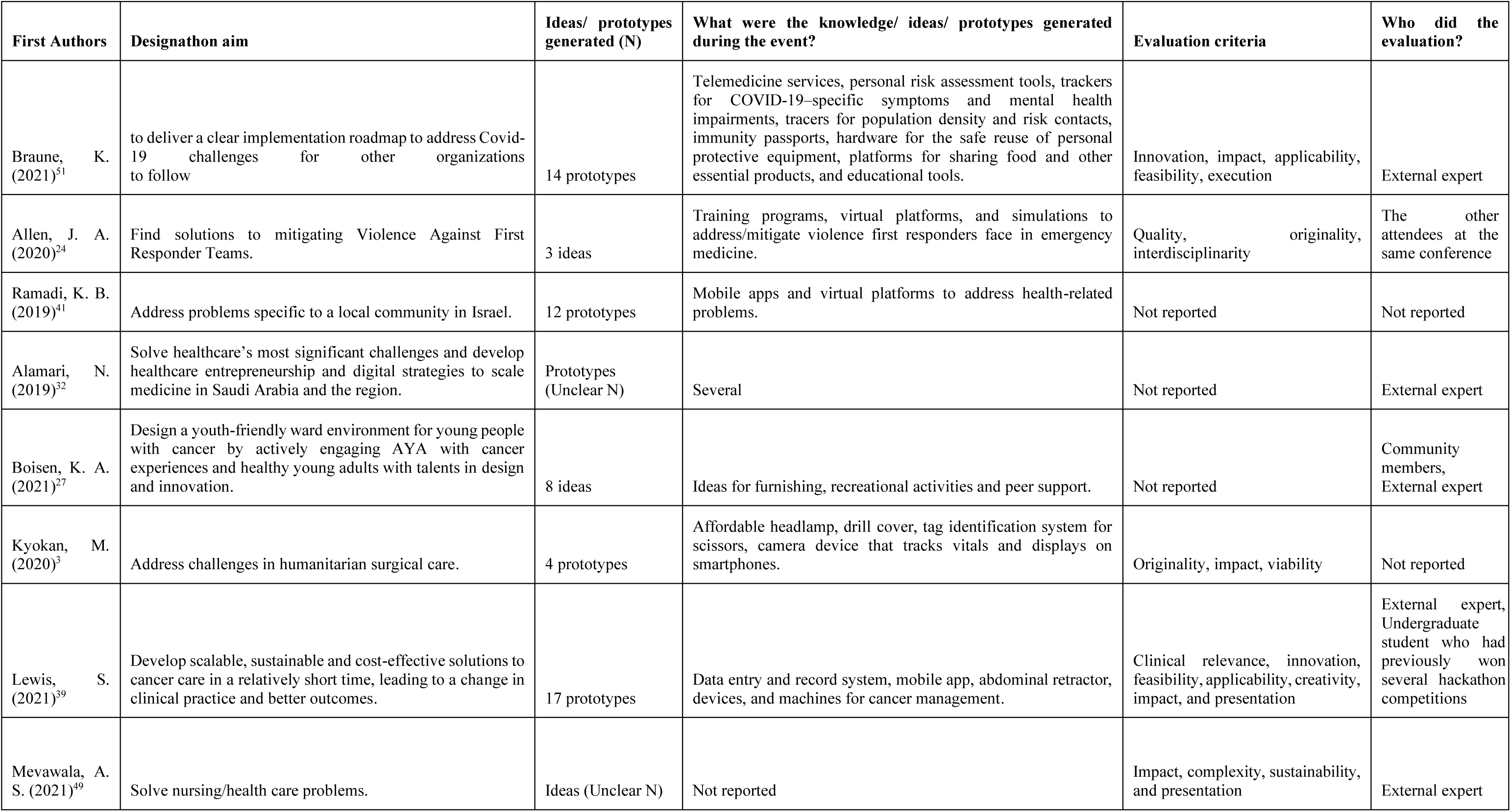
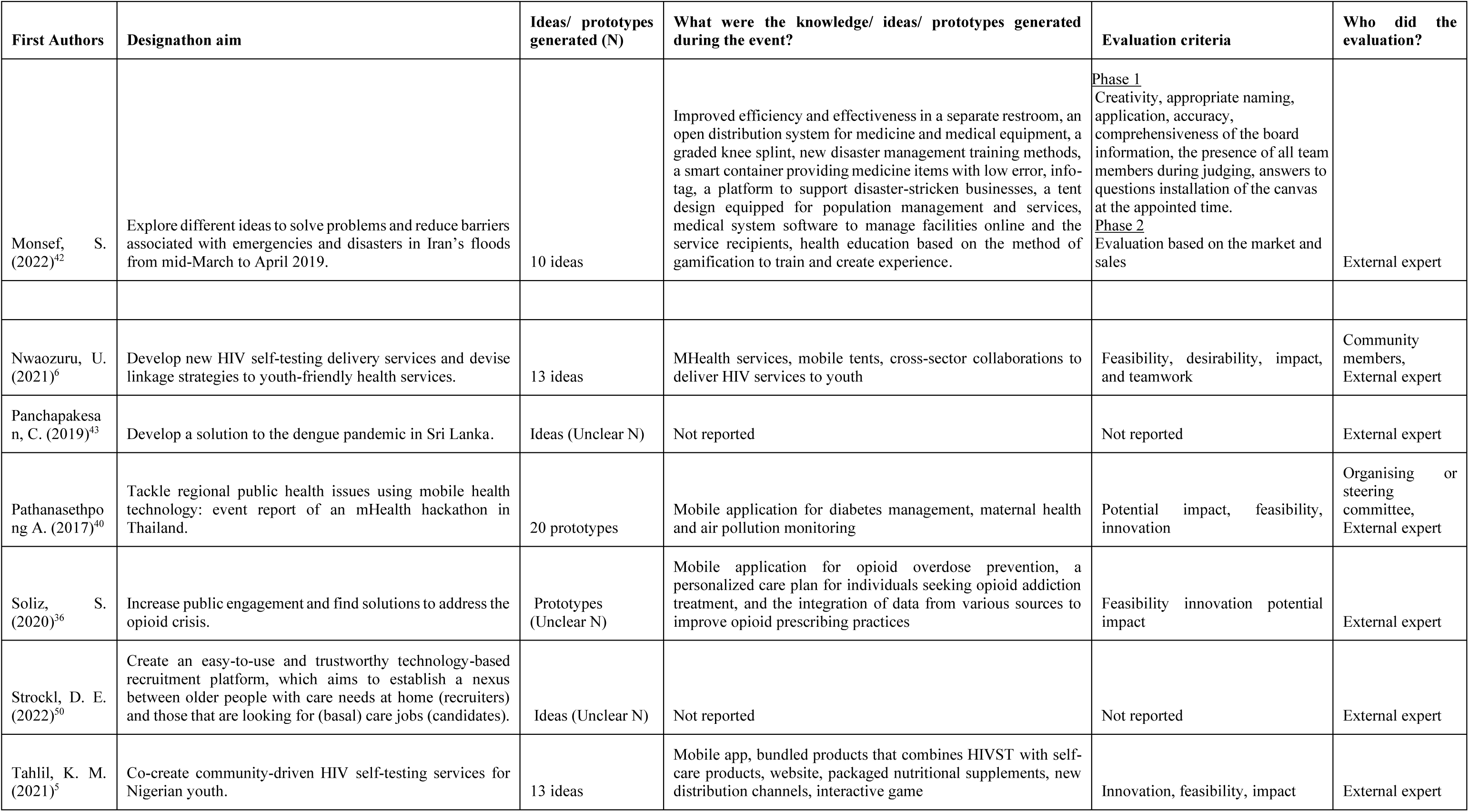
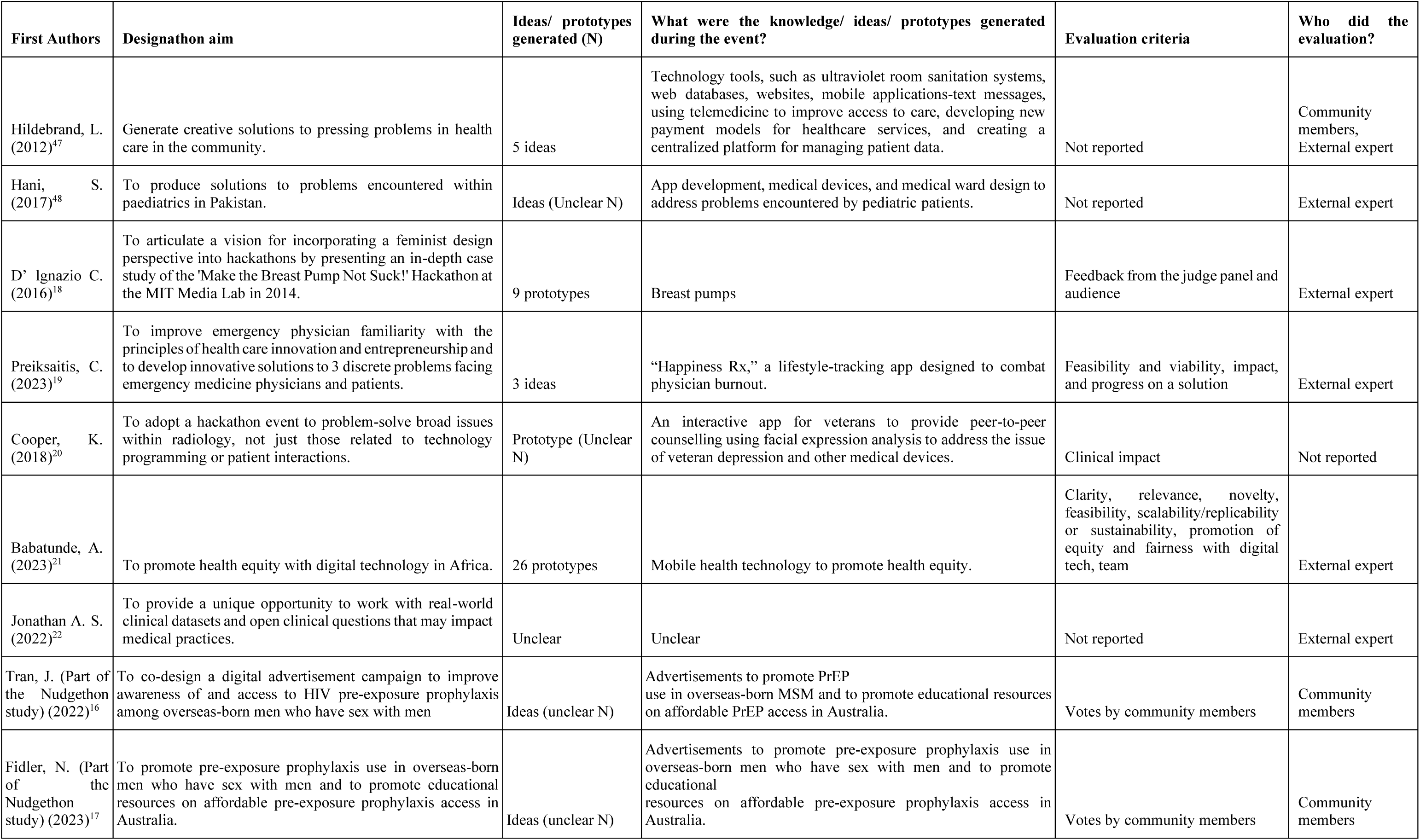
Summary of included studies (N=38)

### Implementation

In 11 studies (29%), ideas or prototypes generated during designathons were further implemented within 12 months,^6, 17, 27, 29, 35, 39, 41–43, 47, 49^ with only four studies (11%) reported testing those ideas or prototypes in the real world via an online survey and research study (Table 3).^6, 17, 29, 35^

### Facilitators of a successful designathon

Twenty-eight (74%) studies reported facilitators of a successful designathon.^3, 5, 6, 16, 22–24, 26–28, 30, 33, 35, 36, 38–40, 42, 45, 47–51^ The most common facilitator reported was including diverse interdisciplinary participants (n=19, 50%),^3, 5, 16, 22, 24, 26, 27, 30, 33, 35, 36, 38–40, 45, 47, 48, 50, 51^ followed by high-quality mentoring/coaching for designathon participants (n=13, 34%),^3, 5, 16, 26, 33, 35, 36, 38–40, 42, 49, 51^ and identifying participants’ with contextual knowledge and/or familiarity with the target problem (n=8, 21%).^3, 26, 31, 33, 35, 39, 40, 50^ Other facilitators reported included clear communication plans (n=7, 18%),^5, 22, 23, 28, 33, 36, 45^ conducive workshop environment (n=7, 18%),^6, 22, 27, 35, 38, 39, 45^ team-building activities (n=6, 16%),^24, 27, 28, 39, 40, 51^ partnership with organisations and stakeholders (n=4, 11%),^6, 32, 38, 41^ virtual workshop options (n=3, 8%),^16, 26, 51^ and the collocation of designathons within larger events (n=2, 5%).^24, 49^

### Barriers to organizing a designathon

Eighteen studies (47.4%) highlighted various barriers encountered during organization designathon events.^3, 5, 16, 20–22, 24, 26, 29, 33, 34, 36, 39, 41, 43–45, 51^ These barriers can be classified into three main themes: 1) insufficient planning and implementation the designathon events (n=13, 34.2%);^3, 5, 21, 24, 26, 29, 34, 36, 39, 43–45, 51^ 2) difficulties in recruiting diverse participants (n=8, 20.1%);^3, 20, 26, 29, 33, 36, 39, 41^ and 3) limited onsite mentorship (n=3, 7.9%).^3, 39, 51, 52^ The engagement barriers primarily revolved around recruiting interdisciplinary participants, balancing each profession across teams, and effectively collaborating with team members holding divergent perspectives. In the implementation aspect, the main barriers included securing funding, technology-related issues during the event, and insufficient time for event planning, implementing and post-event follow-up or allowing the participants to carry on their ideas forward. For example, while organizing designathon events within existing conferences could be convenient, careful planning in advance was necessary to ensure sufficient time allocation. For participants who typically had demanding schedules (e.g., medical professionals, policymakers), organizing an intense in-person eventlasting for two or three days would be difficult. The challenges of providing mentorship of good quality included unclear communication or limited and unstructured post-event support, which affects the long-term sustainability of projects.

### Strengths of using a designathon

Thirty-one studies (82%) explicitly discussed the strengths of the designathon approach.^3, 5, 6, 18–24, 26–33, 35–45, 47–51^ The strengths emerged into 10 themes as follows: 1) engaging an interdisciplinary set of participants (61%, n= 23);^3, 5, 24, 26, 28, 30, 31, 33, 35–39, 41–45, 47, 49–51^ 2) encouraging creativity and innovation (61%, n=23);^3, 5, 6, 23, 26–28, 30–33, 35, 36, 38, 39, 41–44, 47, 49, 51^ 3) creating ways for end-users to lead in the development (47%, n=18);^3, 5, 6, 19, 26–29, 31, 35, 36, 38–42, 47, 50, 51^ 4) networking, connections, and collaborations (47%, n=18);^3, 5, 6, 24, 26, 27, 29–31, 35, 36, 41–45, 49, 51^ 5) being faster (13%, n=5);^5, 28, 36, 47, 51^ and 6) being cheaper (5%, n=2) as strengths.^28, 41^ Other strengths that were included in individual articles were that the designathon was 7) free, open for all to participate, 8) accelerated the development of ideas, 9) engaged the broader society in a topical area, and 10) increased the likelihood of community engagement for a proposed solution.

### Limitations of using a designathon

Twenty-three studies (61%) explicitly discussed the limitations of the designathon approach. The limitations emerged into seven themes as follows: 1) the generalizability of findings generated from designathons (24%, n=9);^5, 6, 19, 25, 30, 36, 41, 43, 44^ 2) the intensive time commitment required to participate (18%, n=7);^5, 18, 26, 36, 41, 43, 49^ 3) challenges associated with assessing impact (16%, n=6);^5, 31, 33, 45, 47, 49^ 4) location if the event is held in-person (13%, n=5);^25, 31, 33, 38, 45^ 5) participant stigma or sampling bias (8% respectively, n=3);^6, 39, 41^ 6) cost (5%, n=2);^23, 49^ and 7) concerns regarding long-term potential, such as low idea feasibility, or insufficient support or opportunities to sustain initial idea investments (17%, n=4).^3, 6, 32, 33^ Other limitations that were identified in individual articles included concerns that designathons encourage competition rather than collaboration, focus on earning a prize rather than facilitating true impact, ideas are not always feasible, and participants may not be knowledgeable of the communities or settings for which they are developing interventions or prototypes, limiting the applicability of ideas generated during designathons.

## Discussion

This systematic review comprehensively synthesised data from designathons for health. Our review highlights key characteristics of designathons in health research in terms of effectiveness (i.e., engagement, outputs, and implementation), resources, and implementation-related factors (i.e., facilitators, barriers, strengths, and limitations). We found that designathons were an adaptable approach accommodating a wide range of participants and had versatile delivery modes (i.e., in-person, online and hybrid) and time frames. This approach also offered flexibility in terms of participant scalability and required resources. By engaging interdisciplinary participants and end users, designathons encouraged creativity and innovation.

Our data suggest that designathons can spark creativity in partnership with end users. Designathons provide a supportive environment that facilitates intensive collaboration between interdisciplinary participants and end users, thereby enabling the development of user-led innovations. Additionally, the guidance and support from mentors enhanced the workshop experience and the quality of solutions generated. The participatory nature of designathons can enhance the likelihood of implementation success and prolong the solution sustainability since the solutions align with the needs of the end-users. In addition to their ability to generate effective solutions, designathons have transformative potential due to their adaptability in terms of modes of delivery and required resources. Designathons have been successfully implemented in various settings, ranging from local community organisations,^29^ within conferences^24^ to international platforms.^34, 51^ The diversity of ideas and prototypes generated during designathons emphasises their adaptability since they can be tailored to specific contexts and health challenges. Designathons are inherently action-oriented and are flexible to adapt to meet constantly changing real-world settings and constraints, including modes of delivery and cost. For instance, designathons successfully transitioned to online formats during the Covid-19 pandemic,^16, 17, 51^ which expanded stakeholders’ involvement and reduced organising costs.

While there are several advantages to using designathons to improve health outcomes, previous literature highlighted notable challenges that may limit their utility. First, several studies, for example, pointed out concerns regarding the generalizability of ideas resulting from designathons.^5, 6, 19, 25, 30, 36, 41, 43, 44^ Designathon participants are often tasked with identifying solutions to specific problems in a designated context or population. As such, finalist ideas may not always be generalizable to a wider population. Still, they enable out-of-the-box thinking that could spark new ideas and approaches to solving persistent problems. This suggests the importance of ongoing evaluation of prototypes and ideas for wider applications and their potential for scale-up. Second, a key critique of many designations has been the lack of further development of ideas and/or prototypes after the conclusion of the formal event. This may be due to the heavy focus on prizes or monetary incentives for participation, which encourages participants to emphasise the short-term rather than longer-term impact. To facilitate greater attention to impact and longer-term planning, ongoing collaboration between organisers, sponsors, and participants is needed. Organisers may have to implement programs and practices that support teams in idea implementation or prototype scale-up. Several designathons have provided prize money explicitly to support implementation or research to assess effectiveness. Third, Costs were identified as both a potential advantage and disadvantage, but few studies provided detailed cost information. Further research on the cost-effectiveness of designathons is needed.

Moving forward, there is a need for a practical guide that establishes a shared understanding of applications of designathons in health contexts, outlining core components and the process involved (i.e., plan, implementation, and evaluation). Guidance, tips, and lessons learned from previous designathons should be incorporated into the guide to ensure consistent reporting and assist when comparing between studies. Additionally, further studies on designathons in health should be more comprehensive and robust evaluation plans focusing on long-term outcomes. Key metrics for evaluation should include testing prototypes with end users and evaluating medical or public health outcomes. External evaluations of designathons are necessary to ensure rigor.

Our systematic review has several strengths. First, all peer-reviewed publications from different regions and publications from a complementary open call were comprehensively and systematically reviewed. Second, our review provided a comprehensive understanding of designathons in the context of health since we included evidence from reports, and qualitative and quantitative studies. Therefore, our review offered relevant data to the designathon in health research practical guide developed for World Health Organization and the Special Programme for Research and Training in Tropical Diseases (WHO/TDR). Lastly, we identified knowledge gaps and highlighted areas where future designathons could improve.

Our study has several limitations. First, there were limited data on the cost-effectiveness, further development, and implementation of prototypes following the designathon events. Second, as we restricted the studies to English language publications, we may have missed publications in other languages. Third, some designathon reports may not be published in peer-reviewed journals due to the nature of designathons, which focus on developing ideas for a particular problem rather than generating scientific-based evidence. We mitigated this limitation by including data from the crowdsourced open call for designathons to identify any reports we might miss.

In conclusion, designathons emerge as a promising and effective tool to develop innovative and impactful solutions to address complex health challenges in various settings due to their ability to facilitate collaboration between the community and experts from diverse fields. Given its adaptability (i.e., budget and mode of delivery) and ability to engage a variety of participants’ professions, and end-users, designathons can be implemented in a wide range of circumstances to address various health issues. By harnessing the strengths of designathons, we can unleash new opportunities and empower communities to significantly impact healthcare outcomes. Future work can explore the cost-effectiveness and implementation of designathons to promote diversity and inclusion in health intervention development and implementation.

## Contributors statement

JT and JJO conceived the idea. JC did the search strategy. WT, EC, TR, CL, AB, AO and UN did the screening and extracted the data. All authors contributed to the first draft of the manuscript. WT and EC assessed the risk of bias. All authors contributed to the interpretation of the results. WT did subsequent edits of the manuscript. All authors had final responsibility for the decision to submit for publication.

## Data Availability

All data produced in the present study are available upon reasonable request to the authors

