## Supplementary file for "Designathons in Health Research: A Global Systematic Review"

**Supplementary Material**
**Table S1. Search Strategy Report:**
Date: 11/29/2022
Database: PubMed

| Set # | |
|---|---|
| 1 | Designathons[tiab] OR Designathon[tiab] OR “design a thon”[tiab] OR “design a thons”[tiab] OR design-a-thon[tiab] OR design-a-thons[tiab] OR “design sprint”[tiab] OR “design sprints”[tiab] OR "agile design"[tiab] OR “agile method”[tiab] OR “agile methods”[tiab] OR “agile methodology”[tiab] OR “agile methodologies”[tiab] OR “agile project management”[tiab] OR “scrum framework”[tiab] OR (agile[tiab] AND scrum[tiab]) |
| 2 | ("User-Centered Design"[Mesh] OR Co-create[tiab] OR co-created[tiab] OR co-creation[tiab] OR co-design*[tiab] OR “user-centered design”[tiab] OR “participatory design”[tiab] OR “design thinking”[tiab] OR "Rapid prototyping"[tiab] OR "rapid design"[tiab]) AND (session[tiab] OR sessions[tiab] OR event[tiab] OR events[tiab] OR workshop[tiab] OR workshops[tiab]) |
| 3 | Hackathons[tiab] OR Hackathon[tiab] OR “hack a thon”[tiab] OR “hack a thons”[tiab] OR hack-a-thon[tiab] OR hack-a-thons[tiab] OR Createathons[tiab] OR Createathon[tiab] OR “create a thon”[tiab] OR “create a thons”[tiab] OR create-a-thon[tiab] OR create-a-thons[tiab] OR Makeathons[tiab] OR Makeathon[tiab] OR “make a thon”[tiab] OR “make a thons”[tiab] OR make-a-thon[tiab] OR make-a-thons[tiab] OR Datathons[tiab] OR Datathon[tiab] OR “data a thon”[tiab] OR “data a thons”[tiab] OR data-a-thon[tiab] OR data-a-thons[tiab] OR Mapathons[tiab] OR Mapathon[tiab] OR “map a thon”[tiab] OR “map a thons”[tiab] OR map-a-thon[tiab] OR map-a-thons[tiab] OR Grantathons[tiab] OR Grantathon[tiab] OR “grant a thon”[tiab] OR “grant a thons”[tiab] OR grant-a-thon[tiab] OR grant-a-thons[tiab] OR “hack day”[tiab] OR “hack days”[tiab] OR hackfest[tiab] OR hackfests[tiab] OR codefest[tiab] OR codefests[tiab] OR “hacking marathon”[tiab] OR “hacking marathons”[tiab] |
| 4 | (("interprofessional relations"[mesh] OR "patient care team"[mesh] OR "Community Participation"[Mesh] OR interprofessional[tiab] OR interdisciplinary[tiab] OR multidisciplinary[tiab] OR community[tiab] OR communities[tiab])) AND ((("Cooperative Behavior"[Mesh] OR "patient care team"[mesh] OR "group processes"[mesh] OR Co-create[tiab] OR co-created[tiab] OR co-creation[tiab] OR co-design*[tiab] OR Teamwork[tiab] OR team[tiab] OR teams[tiab] OR "cooperative behavior"[tiab] OR collaborat*[tiab] OR participatory[tiab])) AND ((sprint[tiab] OR sprints[tiab] OR event[tiab] OR events[tiab] OR session[tiab] OR sessions[tiab]) AND ("Diffusion of Innovation"[mesh] OR innovation[tiab] OR innovations[tiab] OR innovate[tiab] OR innovates[tiab] OR innovating[tiab] OR innovated[tiab] OR prototyp*[tiab] OR ideate[tiab] OR ideates[tiab] OR ideated[tiab] ))) |
| 5 | #1 OR #2 OR #3 OR #4 |

Database:  Scopus

| Set # | |
|---|---|
| 1 | TITLE-ABS(Designathons OR Designathon OR “design a thon” OR “design a thons” OR design-a-thon OR design-a-thons OR “design sprint” OR “design sprints” OR "agile design" OR “agile method” OR “agile methods” OR “agile methodology” OR “agile methodologies” OR “agile project management” OR “scrum framework” OR (agile AND scrum)) |
| 2 | TITLE-ABS((Co-create OR co-created OR co-creation OR co-design* OR “user-centered design” OR “participatory design” OR “design thinking” OR "Rapid prototyping" OR "rapid design") AND (session OR sessions OR event OR events OR workshop OR workshops)) |
| 3 | TITLE-ABS(Hackathons OR Hackathon OR “hack a thon” OR “hack a thons” OR hack-a-thon OR hack-a-thons OR Createathons OR Createathon OR “create a thon” OR “create a thons” OR create-a-thon OR create-a-thons OR Makeathons OR Makeathon OR “make a thon” OR “make a thons” OR make-a-thon OR make-a-thons OR Datathons OR Datathon OR “data a thon” OR “data a thons” OR data-a-thon OR data-a-thons OR Mapathons OR Mapathon OR “map a thon” OR “map a thons” OR map-a-thon OR map-a-thons OR Grantathons OR Grantathon OR “grant a thon” OR “grant a thons” OR grant-a-thon OR grant-a-thons OR “hack day” OR “hack days” OR hackfest OR hackfests OR codefest OR codefests OR “hacking marathon” OR “hacking marathons”) |
| 4 | TITLE-ABS((interprofessional OR interdisciplinary OR multidisciplinary OR community OR communities) AND (Co-create OR co-created OR co-creation OR co-design* OR Teamwork OR team OR teams OR "cooperative behavior" OR collaborat* OR participatory) AND (sprint OR sprints OR event OR events OR session OR sessions) AND (innovation OR innovations OR innovate OR innovates OR innovating OR innovated OR prototyp* OR ideate OR ideates OR ideated)) |
| 5 | #1 OR #2 OR #3 OR #4 |
| 6 | TITLE-ABS (health OR healthcare OR medicine OR medical OR wellness OR wellbeing OR Pharmacy OR Pharmaceutical OR patient OR patients) |
| 7 | #5 AND #6 |

Database:  Embase

| Set # | |
|---|---|
| 1 | Designathons:ti,ab,kw OR Designathon:ti,ab,kw OR ‘design a thon’:ti,ab,kw OR ‘design a thons’:ti,ab,kw OR design-a-thon:ti,ab,kw OR design-a-thons:ti,ab,kw OR ‘design sprint’:ti,ab,kw OR ‘design sprints’:ti,ab,kw OR ‘agile design’:ti,ab,kw OR ‘agile method’:ti,ab,kw OR ‘agile methods’:ti,ab,kw OR ‘agile methodology’:ti,ab,kw OR ‘agile methodologies’:ti,ab,kw OR ‘agile project management’:ti,ab,kw OR ‘scrum framework’:ti,ab,kw OR (agile:ti,ab,kw AND scrum:ti,ab,kw) |
| 2 | ('user-centered design'/exp OR Co-create:ti,ab,kw OR co-created:ti,ab,kw OR co-creation:ti,ab,kw OR co-design*:ti,ab,kw OR ‘user-centered design’:ti,ab,kw OR ‘participatory design’:ti,ab,kw OR ‘design thinking’:ti,ab,kw OR ‘Rapid prototyping’:ti,ab,kw OR ‘rapid design’:ti,ab,kw) AND (session:ti,ab,kw OR sessions:ti,ab,kw OR event:ti,ab,kw OR events:ti,ab,kw OR workshop:ti,ab,kw OR workshops:ti,ab,kw) |
| 3 | Hackathons:ti,ab,kw OR Hackathon:ti,ab,kw OR ‘hack a thon’:ti,ab,kw OR ‘hack a thons’:ti,ab,kw OR hack-a-thon:ti,ab,kw OR hack-a-thons:ti,ab,kw OR Createathons:ti,ab,kw OR Createathon:ti,ab,kw OR ‘create a thon’:ti,ab,kw OR ‘create a thons’:ti,ab,kw OR create-a-thon:ti,ab,kw OR create-a-thons:ti,ab,kw OR Makeathons:ti,ab,kw OR Makeathon:ti,ab,kw OR ‘make a thon’:ti,ab,kw OR ‘make a thons’:ti,ab,kw OR make-a-thon:ti,ab,kw OR make-a-thons:ti,ab,kw OR Datathons:ti,ab,kw OR Datathon:ti,ab,kw OR ‘data a thon’:ti,ab,kw OR ‘data a thons’:ti,ab,kw OR data-a-thon:ti,ab,kw OR data-a-thons:ti,ab,kw OR Mapathons:ti,ab,kw OR Mapathon:ti,ab,kw OR ‘map a thon’:ti,ab,kw OR ‘map a thons’:ti,ab,kw OR map-a-thon:ti,ab,kw OR map-a-thons:ti,ab,kw OR Grantathons:ti,ab,kw OR Grantathon:ti,ab,kw OR ‘grant a thon’:ti,ab,kw OR ‘grant a thons’:ti,ab,kw OR grant-a-thon:ti,ab,kw OR grant-a-thons:ti,ab,kw OR ‘hack day’:ti,ab,kw OR ‘hack days’:ti,ab,kw OR hackfest:ti,ab,kw OR hackfests:ti,ab,kw OR codefest:ti,ab,kw OR codefests:ti,ab,kw OR ‘hacking marathon’:ti,ab,kw OR ‘hacking marathons’:ti,ab,kw |
| 4 | ('collaborative care team'/exp OR 'community participation'/exp OR interprofessional:ti,ab,kw OR interdisciplinary:ti,ab,kw OR multidisciplinary:ti,ab,kw OR community:ti,ab,kw OR communities:ti,ab,kw) AND ('cooperation'/exp OR 'group process'/exp OR Co-create:ti,ab,kw OR co-created:ti,ab,kw OR co-creation:ti,ab,kw OR co-design*:ti,ab,kw OR Teamwork:ti,ab,kw OR team:ti,ab,kw OR teams:ti,ab,kw OR ‘cooperative behavior’:ti,ab,kw OR collaborat*:ti,ab,kw OR participatory:ti,ab,kw) AND (sprint:ti,ab,kw OR sprints:ti,ab,kw OR event:ti,ab,kw OR events:ti,ab,kw OR session:ti,ab,kw OR sessions:ti,ab,kw) AND ('diffusion of innovation'/exp OR innovation:ti,ab,kw OR innovations:ti,ab,kw OR innovate:ti,ab,kw OR innovates:ti,ab,kw OR innovating:ti,ab,kw OR innovated:ti,ab,kw OR prototyp*:ti,ab,kw OR ideate:ti,ab,kw OR ideates:ti,ab,kw OR ideated:ti,ab,kw ) |
| 5 | #1 OR #2 OR #3 OR #4 |
| 6 | #5 AND [embase]/lim NOT ([embase]/lim AND [medline]/lim) |
| 7 | #6 AND ('article'/it OR 'article in press'/it OR 'conference paper'/it OR 'conference review'/it OR 'editorial'/it OR 'erratum'/it OR 'letter'/it OR 'note'/it OR 'review'/it OR 'short survey'/it) |

Database: Cochrane Library

| Set # | |
|---|---|
| 1 | Designathons:ti,ab OR Designathon:ti,ab OR “design a thon”:ti,ab OR “design a thons”:ti,ab OR design-a-thon:ti,ab OR design-a-thons:ti,ab OR “design sprint”:ti,ab OR “design sprints”:ti,ab OR "agile design":ti,ab OR “agile method”:ti,ab OR “agile methods”:ti,ab OR “agile methodology”:ti,ab OR “agile methodologies”:ti,ab OR “agile project management”:ti,ab OR “scrum framework”:ti,ab OR (agile:ti,ab AND scrum:ti,ab) |
| 2 | ([mh "User-Centered Design"] OR Co-create:ti,ab OR co-created:ti,ab OR co-creation:ti,ab OR co-design*:ti,ab OR “user-centered design”:ti,ab OR “participatory design”:ti,ab OR “design thinking”:ti,ab OR "Rapid prototyping":ti,ab OR "rapid design":ti,ab) AND (session:ti,ab OR sessions:ti,ab OR event:ti,ab OR events:ti,ab OR workshop:ti,ab OR workshops:ti,ab) |
| 3 | Hackathons:ti,ab OR Hackathon:ti,ab OR “hack a thon”:ti,ab OR “hack a thons”:ti,ab OR hack-a-thon:ti,ab OR hack-a-thons:ti,ab OR Createathons:ti,ab OR Createathon:ti,ab OR “create a thon”:ti,ab OR “create a thons”:ti,ab OR create-a-thon:ti,ab OR create-a-thons:ti,ab OR Makeathons:ti,ab OR Makeathon:ti,ab OR “make a thon”:ti,ab OR “make a thons”:ti,ab OR make-a-thon:ti,ab OR make-a-thons:ti,ab OR Datathons:ti,ab OR Datathon:ti,ab OR “data a thon”:ti,ab OR “data a thons”:ti,ab OR data-a-thon:ti,ab OR data-a-thons:ti,ab OR Mapathons:ti,ab OR Mapathon:ti,ab OR “map a thon”:ti,ab OR “map a thons”:ti,ab OR map-a-thon:ti,ab OR map-a-thons:ti,ab OR Grantathons:ti,ab OR Grantathon:ti,ab OR “grant a thon”:ti,ab OR “grant a thons”:ti,ab OR grant-a-thon:ti,ab OR grant-a-thons:ti,ab OR “hack day”:ti,ab OR “hack days”:ti,ab OR hackfest:ti,ab OR hackfests:ti,ab OR codefest:ti,ab OR codefests:ti,ab OR “hacking marathon”:ti,ab OR “hacking marathons”:ti,ab |
| 4 | ([mh "interprofessional relations"] OR [mh "patient care team"] OR [mh "Community Participation"] OR interprofessional:ti,ab OR interdisciplinary:ti,ab OR multidisciplinary:ti,ab OR community:ti,ab OR communities:ti,ab) AND ([mh "Cooperative Behavior"] OR [mh "patient care team"] OR [mh "group processes"] OR Co-create:ti,ab OR co-created:ti,ab OR co-creation:ti,ab OR co-design*:ti,ab OR Teamwork:ti,ab OR team:ti,ab OR teams:ti,ab OR "cooperative behavior":ti,ab OR collaborat*:ti,ab OR participatory:ti,ab) AND (sprint:ti,ab OR sprints:ti,ab OR event:ti,ab OR events:ti,ab OR session:ti,ab OR sessions:ti,ab) AND ([mh "Diffusion of Innovation"] OR innovation:ti,ab OR innovations:ti,ab OR innovate:ti,ab OR innovates:ti,ab OR innovating:ti,ab OR innovated:ti,ab OR prototyp*:ti,ab OR ideate:ti,ab OR ideates:ti,ab OR ideated:ti,ab) |
| 5 | #1 OR #2 OR #3 OR #4 |

Database: ClinicalTrials.gov

| Set # | |
|---|---|
| 1 | Designathons OR Designathon OR “design a thon” OR “design a thons” OR design-a-thon OR design-a-thons |
| 2 | (Co-create OR co-created OR co-creation) AND (event OR events OR workshop OR workshops OR session OR sessions) AND (community OR participatory) |
| 3 | Hackathons OR Hackathon OR “hack a thon” OR “hack a thons” OR hack-a-thon OR hack-a-thons |
| 4 | (Teamwork OR cooperative) AND (interprofessional OR interdisciplinary OR multidisciplinary OR business OR engineering) and innovation |
| 5 | #1 OR #2 OR #3 OR #4 |

### Table S2. Risk of bias assessment

| Author | Year | 1. Were respondents' demographic characteristics clearly described? | 2. Was the problem(s) clearly described? | 3. Were evaluation methods clearly described? | 4. Were the result(s) or final product(s) clearly described? | 5. Was the implementation after events clearly described? | 6. Does the report provide takeaway lessons? | Overall appraisal: |
|---|---|---|---|---|---|---|---|---|
| Morales, E. (2018)[1] | 2018 | Yes | Yes | Yes | Yes | No | Yes | Yes |
| Wang, J. K. (2018)[2] | 2018 | Yes | Yes | Yes | Yes | NA | Yes | Yes |
| Mirkovic, J. (2018)[3] | 2021 | Yes | Yes | No | Yes | NA | Yes | Yes |
| Shonkoff, E. T. (2021)[4] | 2022 | Yes | Yes | Yes | No | NA | Yes | Yes |
| Bogomolova, S. (2020)[5] | 2019 | No | Yes | No | Yes | Yes | Yes | Yes |
| Pakpour, N. (2022)[6] | 2017 | Yes | Yes | NA | Yes | NA | Yes | Yes |
| Hope, A. (2019)[7] | 2020 | Yes | Yes | Yes | Yes | No | Yes | Yes |
| Birbeck, N. (2017)[8] | 2020 | Yes | Yes | Yes | Yes | No | Yes | Yes |
| Cardwell, F. S. (2021)[9] | 2020 | Yes | Yes | Yes | Yes | NA | Yes | Yes |
| Li, C.(2020)[10] | 2016 | Yes | Yes | No | No | NA | Yes | Yes |
| Ling, R.(2021)[11] | 2020 | No | Yes | Yes | Yes | NA | Yes | Yes |
| Poncette, A. S. (2020)[12] | 2019 | Yes | Yes | No | No | NA | Yes | Yes |
| Silver, J. K.(2016)[13] | 2015 | No | Yes | No | Yes | No | Yes | Yes |
| Wang, J. K.(2018)[14] | 2020 | No | Yes | Yes | Yes | NA | Yes | Yes |
| Braune, K. (2021)[15] | 2021 | Yes | Yes | Yes | Yes | Yes | Yes | Yes |
| Allen, J. A. (2020)[16] | 2021 | No | Yes | Yes | No | No | Yes | Yes |
| Ramadi, K. B. (2019)[17] | 2022 | Yes | Yes | Yes | Yes | No | Yes | Yes |
| Alamari, N. (2019)[18] | 2021 | Yes | Yes | Yes | Yes | Yes | Yes | Yes |

**Table S2 (cont). Risk of bias assessment**

| Author | Year | 1. Were respondents' demographic characteristics clearly described? | 2. Was the problem(s) clearly described? | 3. Were evaluation methods clearly described? | 4. Were the result(s) or final product(s) clearly described? | 5. Was the implementation after events clearly described? | 6. Does the report provide takeaway lessons? | Overall appraisal: |
|---|---|---|---|---|---|---|---|---|
| Boisen, K. A. (2021)[19] | 2017 | No | Yes | Yes | Yes | NA | Yes | Yes |
| Kyokan, M. (2020)[20] | 2022 | No | Yes | No | No | NA | No | Yes |
| Lewis, S. (2021)[21] | 2021 | Yes | Yes | Yes | Yes | NA | Yes | Yes |
| Mevawala, A. S. (2021)[22] | 2012 | No | Yes | No | Yes | Yes | Yes | Yes |
| Monsef, S. (2022)[23] | 2017 | No | Yes | No | Yes | NA | Yes | Yes |
| Nwaozuru, U. (2021)[24] | 2016 | No | Yes | No | Yes | Yes | Yes | Yes |
| Panchapakesan, C. (2019)[25] | 2023 | No | Yes | Yes | Yes | NA | Yes | Yes |
| Pathanasethpong A. (2017)[26] | 2018 | No | Yes | Yes | Yes | NA | Yes | Yes |
| Soliz, S. (2020)[27] | 2023 | No | Yes | Yes | Yes | NA | Yes | Yes |
| Strockl, D. E. (2022)[28] | 2023 | Yes | Yes | No | Yes | Yes | Yes | Yes |
| Tahlil, K. M. (2021)[29] | 2018 | Yes | Yes | No | Yes | NA | Yes | Yes |
| Hildebrand, L. (2012)[30] | 2020 | Yes | Yes | No | Yes | Yes | Yes | Yes |
| Hani, S. (2017)[31] | 2021 | Yes | Yes | Yes | Yes | NA | Yes | Yes |
| D' lgnazio C. (2016)[32] | 2018 | Yes | Yes | Yes | Yes | NA | Yes | Yes |
| Preiksaitis, C. (2023)[33] | 2021 | No | Yes | Yes | Yes | NA | Yes | Yes |
| Cooper, K. (2018)[34] | 2019 | Yes | Yes | No | Yes | No | Yes | Yes |
| Babatunde, A. (2023)[35] | 2019 | Yes | Yes | No | No | No | Yes | Yes |

**Table S2 (cont). Risk of bias assessment**

| Author | Year | 1. Were respondents' demographic characteristics clearly described? | 2. Was the problem(s) clearly described? | 3. Were evaluation methods clearly described? | 4. Were the result(s) or final product(s) clearly described? | 5. Was the implementation after events clearly described? | 6. Does the report provide takeaway lessons? | Overall appraisal: |
|---|---|---|---|---|---|---|---|---|
| Jonathan A. S. (2022)[36] | 2020 | No | Yes | Yes | Yes | NA | Yes | Yes |
| Tran, J. (Part of the Nudgethon study) (2022)[37] | 2022 | Yes | Yes | No | No | NA | Yes | Yes |
| Fidler, N. (Part of the Nudgethon study) (2023)[38] | 2022 | Yes | Yes | No | Yes | Yes | Yes | Yes |

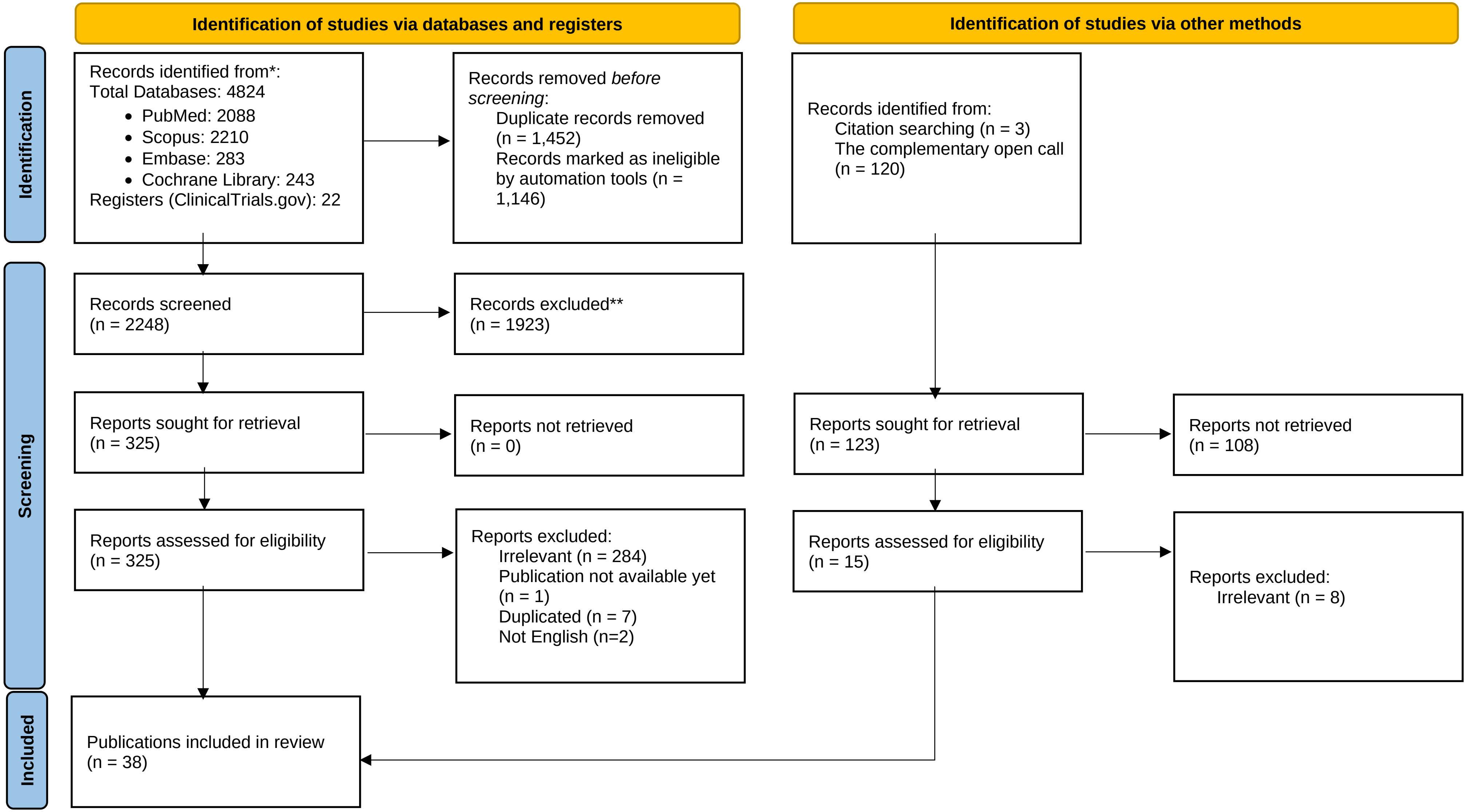


*Consider, if feasible to do so, reporting the number of records identified from each database or register searched (rather than the total number across all databases/registers).
**If automation tools were used, indicate how many records were excluded by a human and how many were excluded by automation tools.

**Figure S1. PRISMA flowchart of the study selection proces**
